# Factors Associated with Malaria Infection in Under Five Children-Zambia, 2018. A Secondary Analysis of the Malaria Indicator Survey, 2018

**DOI:** 10.1101/2023.09.05.23295063

**Authors:** Lindiwe Tembo, Cephas Sialubanje, Martha Malasa, Elizabeth Susan Heilmann, Nyambe Sinyange, Mukumbuta Nawa

## Abstract

**Introduction:** Malaria is a public health concern in Zambia. In 2018, Zambia reported a 9% malaria prevalence in under-five children, with some provinces reporting above 20%. Factors still driving malaria infection among the country’s under-five children must be investigated. Factors associated with malaria infection among under-five children were investigated.

**Methods:** This was a secondary analysis of the cross-sectional study for the Zambia Malaria Indicator Survey 2018 (MIS). Multistage sampling was used in the malaria indicator survey. All children aged 6 to 59 months who received malaria rapid diagnostic tests (RDT) in the data set were considered for the study. Malaria infection: Tested positive or negative for malaria RDT. Sample weights and multivariable logistic regression were used. Backward stepwise regression was used to determine the best model, and the Akaike information criterion was used to select the best model that best fits the data. The Odds ratio measured the association at a 95% confidence level.

**Results:** A total of 2400 children were analysed. 24.3% (583) tested positive. The median age was 32 (interquartile range (IQR:8-46)) months. Males were 52% (1,249). The odds of malaria infection increased as the child’s age in months increased (aOR=1.004, 95% CI: 1.003, 1.005). Children in the rural had higher odds of malaria than urban children (aOR=1.11,95% CI: 1.05, 1.17). The odds of malaria in children in Copperbelt, Luapula, Muchinga and North-Western Provinces were higher than in children in Central Province (p-value <0.05). Children whose houses did not receive IRS had increased odds of malaria compared to children whose houses received IRS (aOR=1.05, 95%CI: 1.01, 1.09). Sex was not statistically significant.

**Conclusion:** An increase in age, living in the rural and northern parts of Zambia was associated with malaria infection. Increasing malaria prevention and control measures for older children, rural communities, and the northern parts of Zambia may help reduce malaria prevalence.

**Strengths and Limitations:**

1. The study used a large dataset of the Malaria Indicator Survey 2018 that was powered to be nationally representative of Zambia, urban and rural strata, and provincial levels.
2. The study used a complex data analysis considering household weights, census enumeration areas and provinces, addressing intra-cluster correlations.
3. The study addressed confounding using multivariable regressing and determined the best-fit model using Akaike Information Criteria (AIC) in a backward stepwise regressing.
4. The study’s main limitation is that it is a survey that simultaneously assessed the outcome and exposure variables, eliciting associations only and not causality.

## INTRODUCTION

Malaria is a public health concern with the most burden in sub-Saharan Africa (1). It is a parasitic infection transmitted by a female *Anopheles* mosquito (1). Globally, malaria infection reduced from about 238 million cases in 2000 to 217 million cases in 2014, with an increase to 229 million cases in 2019 (2).

Globally, as of 2019, 3.2 billion people were at risk of being infected with malaria (3) and about 229 million cases were reported in 2019, leading to 405,000 deaths. The under-five children account for 67% of the deaths reported (4,5).

Significantly, most malaria morbidity (93%) and mortality (91%) occur in sub-Saharan Africa. Zambia contributes 2% of the world’s malaria burden and 5.2% in East and Southern Africa (6). It recorded a 26.8% reduction in malaria cases from 202 to 148 cases per 1000 population from 2016 to 2018, with its districts reporting incidences of 50-500 per 1000 population at risk between 2016 and 2019 (7). Mortality rates reduced by 5.4% from 0.47 to 0.44 per 1000 population between 2016 to 2019. Zambia’s malaria prevalence in children under five has reduced from 22% in 2006 to 9% in 2018 (7). However, in 2020, Zambia recorded a 30 - 50% increase in malaria cases, deaths, and test positivity rates compared to 2018 and 2019(8).

Children under five years are one of the most vulnerable groups. They are highly susceptible to infection in areas with high malaria transmission causing one death every two minutes in Africa and one death in every five under-five children in Zambia (9). Malaria is Zambia’s second top cause of death, recording 12% of the total deaths. Under-five children contribute more than 50% of these deaths (10). Malaria in under-five children indicates the malaria burden in an area or country (11).

Therefore, the Government of the Republic of Zambia (GRZ), through the National Malaria Elimination Program (NMEP), PMI, Global Fund, and other partners, have set a goal to eliminate malaria in local communities by implementing various interventions and include promotion and distribution of ITN for pregnant women and under-five children, use of artemisinin combination therapy (ACT) for treatment, IRS, larviciding and intermittent prophylactic therapy in pregnant women (IPTp) among others (12). Despite implementing these interventions, malaria remains a health challenge in under-five children. This study sought to investigate factors still driving malaria among under-five children despite efforts to eliminate the disease in Zambia.

## METHODOLOGY

### Study Site and Setting

The study was conducted in Zambia. Zambia is a plateau located in the Southern part of Africa. The total area is 752,614-kilometre square. It has a tropical climate with an average rainfall of 500 mm in the southern part to 1400 mm in the northern parts of Zambia annually. The average temperatures are 20 ° C in the winter and 30 ° C in the summer and rainy seasons. Zambia has a population of over 19 million as of 2021 (13).

### Study Population

This study analysed data from all ten provinces of Zambia, and the study population are children from 6 to 59 months of age. This study population is all children from six to 59 months of age tested for malaria using the Rapid Diagnostic Test (RDT) for malaria during the MIS, 2018.

### Study Design

This was the analysis of the cross-sectional dataset, the Zambia Malaria Indicator Survey, 2018.

The 2018 MIS collected data from mid-April to late May of 2018. This time corresponds to the last part of the rains in Zambia and peak malaria transmission. This study was a cross-sectional survey designed as a representative probability sample to produce the provincial and country estimates and separate rural and urban residences. The country is divided into ten provinces, and they are divided into districts. The districts are divided into census supervisory areas (CSA), which are further divided into standard enumeration areas (SEA). The number of households in a SEA was collected from the 2010 population census. The sampling frame was the list of SEAs. A total of 4,475 households were planned to be selected from about 179 SEAs, and the number of children to receive malaria testing was under six years and ten years in the Western province (7).

### Inclusion and Exclusion Criteria

#### Inclusion Criteria

The study included all the children from 6 to 59 months of age in the 2018 Malaria Indicator Survey who received a malaria rapid diagnostic test (RDT).

#### Exclusion Criteria

Exclusion criteria included children aged 6 to 59 months who did not receive a malaria diagnostic test (RDT).

#### Data Collection Tool and Techniques

Data was collected from the MIS 2018 data set and cleaned using the data extraction tool. The USAID collected data three months before the MIS started; data for January 2018 was merged with the MIS 2018 data set and analysed.

### Malaria Indicator Survey

Zambia National Malaria Indicator Survey (MIS) 2018 is a malaria-focused household survey undertaken to benchmark the progress of malaria burden reduction and coverage of key malaria interventions such as ITN and IRS in Zambia. The MIS is important in assessing and monitoring changes in parasite prevalence in under-five children.

### CHIRPS

Climate hazard infrared precipitation with stations (CHIRPS) has developed techniques for producing rainfall maps. Average rainfall estimates were derived from the satellite data. It delivers complete, reliable, up-to-date datasets on rainfall trends. This information was useful in relating to malaria infection in various regions of Zambia.

### Entomological Malaria Dataset

This dataset contains data on mosquito vectors captured and analysed for specie identification, densities and sporozoite rate. The data set also includes morphology, physiology, anatomy, behaviour, distribution, and its relationship to the environment. However, in this study, we only had access to vector distribution in the provinces of Zambia. These datasets were merged with the MIS 2018 dataset.

### Sample Size Determination

The sample size was determined by using the Leslie Kish sample size determination(14). This formula has design effects and accounts for stratification and weights for multistage sampling. The malaria prevalence was 9% in Zambia in 2018. Precision set at 5%, the design effect for MIS 2018 was 2.0, and the non-response was 20%. The desired sample size was calculated as 302. The data set had a total of 2400 observations. The study has enough power.

### Data Analysis

R-Studio 4.04 was used to conduct data analysis and visualisation(15). Descriptive data were summarised using frequency tables. The data were not normally distributed. Wilcoxon-rank sum tests were used to measure differences in the median.

Bivariate logistic regression was used to determine the association between the dependent and independent variables. Multivariable logistic regression was used to predict the relationship between dependent and independent variables and control for confounding. Only variables with p-value < 0.20 were added to the multivariable logistic regression. Sample weights were used to calculate population estimates for the survey design. Odds ratios were used to measure the association between the dependent and independent variables at a 95% confidence level. A p-value less than 0.05 was considered significant.

### Ethical Consideration

ERES Converge Board cleared this research study. The Zambia National Health Research Authority then authorised it. Further approval was sought from the Permanent Secretary of the Ministry of Health and Malaria Elimination Control Centre to obtain the data set. We also secured the data set in a secured laptop with a password.

## RESULTS

### Participants of the Study

The Malaria Indicator dataset had 2,768 children between the ages of <1 month to 59 months. There were 259 observations below the age of 6 months, 40 had missing RDT results, and 69 had missing weights. In this study, the data for 2400 children who were between the age of 6 to 59 months were analysed, as shown in Fig 1.

**Figure 1:**
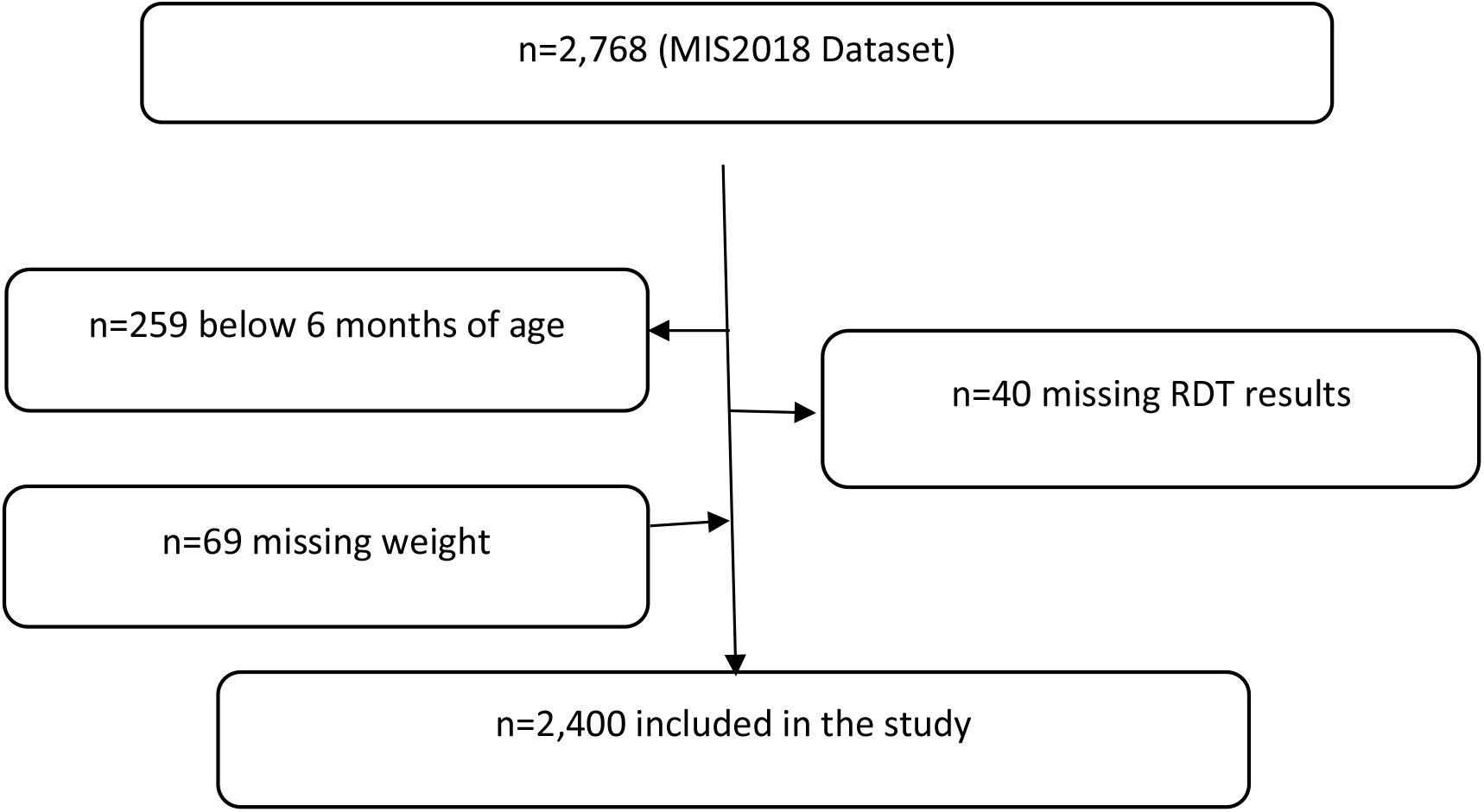
Flow Chart on Inclusion and Exclusions.

### Demographic and Exposure Characteristics of Malaria

Table 1 shows the characteristics of study participants by malaria status. The median age was 32 (interquartile range [IQR]:18-46) months. Males were 52% (1,249), and females were 48% (1,151). National malaria parasite prevalence (RDT) was 24.3% (583). The province with the lowest prevalence was Lusaka and Southern, reporting 0.3%, and Central province reporting 2.7%. The province with the highest prevalence was Luapula, with 32.8%, followed by Western at 15.4% and Eastern at 4.8%. The prevalence of malaria in rural areas was 28.2% (562), and in urban was 5.2% (21). The total prevalence was 28.2% (583).

Chi-square analysis showed that children testing malaria positive and malaria negative were statistically different in age, sex, living in rural/urban, province, wealth quantile, toilet, wall, roof, ceiling, window, indoor residual spray (IRS), cluster IRS coverage, eaves, ill with a fever last two weeks, sleep in ITN last night, haemoglobin level, *Anopheles Gambie, Anopheles Funestus*, rainfall and household energy.

### The Factors Associated with Malaria Infection in Under-Five Children

Table 2 shows the full model, Table 3 the model fitness using the Akaike Information Criteria (AIC), and Table 4 shows the reduced model after performing model fitness. The odds of malaria infection increased as children’s age in months increased (aOR=1.003, 95% CI: 1.002, 1.01). Children living in rural areas had higher odds of malaria infection than those in urban areas (aOR=1.12, 95%CI:1.06,1.18). Compared to the Central province, the odds of malaria in children living in Copperbelt, Luapula, Muchinga and North-western provinces had higher odds of malaria (aOR= 1.14, 1.03, 1.28, aOR = 1.35, 95%CI: 1.20, 1.52; aOR = 1.25, 95%CI: 1.08, 1.44, aOR = 1.34, 95%CI: 1.13, 1.58).

Compared to children from the lowest wealth quintile, children who came from homes whose wealth quintile was middle and middle high were 88% and 91% less likely to be infected with malaria (95% CI: 82%, 0.94, 95% CI: 82%, 99%). Children whose haemoglobin level was less than 6.9g/dl had increased odds of malaria compared to children whose haemoglobin level was more than 6.9g/dl (aOR=1.20, 95%CI: 1.02, 1.42).

Children whose house was not sprayed against mosquitoes had higher odds of malaria infection (aOR = 1.05, 95% CI: 1.01,1.10). Compared to children whose houses had completely closed eaves, children whose houses had partially closed eaves were 95% less likely to have malaria (95% CI: 91%, 99%). Compared to children whose houses had a completely sealed ceiling, compared to children whose houses were partially sealed were 89% less likely to have malaria (95% CI: 0.80, 0.99). Compared to children whose houses had windows and airbricks completely sealed with screens, children whose houses had windows and airbricks with holes had higher odds of malaria infection (aOR= 1.09, 95% CI: 1.02, 1.17)

Children whose household water source was a we1l, stream or river had higher odds of malaria infection than children whose households had piped water (aOR=1.06, 95%CI: 1.01, 1.11). Household head education level, type of walls, type of toilet, fuel used for the household, time child got out of bed, and average rainfall was not statistically significant.

## DISCUSSION

An increase in the age of children was associated with malaria infection. The older children may not receive protection and care from their mothers. These children may no longer have maternal immunity and not sleep close to their mothers; hence may no longer benefit from ITN protection. A study conducted in Malawi and Nigeria showed that older children had higher odds of malaria infection (16,17).

Children from rural areas had increased odds of malaria infection compared to children in urban areas of Zambia. This may be because of poor malaria interventions implemented in rural areas. The urban population may be privileged to access other protective factors such as easy access to ITNs, mosquito repellants, proper housing structures, and access to health care. This is similar to a study conducted in Nigeria that reviewed that rural populations had higher odds of malaria than the urban population due to inadequate malaria interventions and that the rural communities might not be able to afford to purchase malaria intervention items such as ITNs (17).

Children living in Copperbelt, Luapula, Muchinga, and North-western Provinces had higher odds of malaria. Environmental factors such as rainfall partly explain the high number of malaria infections in these provinces. These provinces are characterised by high rainfall of over 800ml/m. Rainwater is collected in ditches, depressions, streams, and rivers making many areas suitable for bleeding. These provinces also have many water bodies, such as rivers, lakes, dams, and streams. The water bodies are freshwater bodies and are surrounded by swampy areas. This environment has been conducive to breeding Anopheles mosquitoes, hence the high transmission of malaria parasites. Rainfall also increases the humidity, which prolongs adult mosquito life. During the rainy season, the temperatures are also favourable for mosquito breeding (18). The annual average rainfall is about 960mm, with the northern part receiving up to 1500mm and the southwest receiving about 660mm. The rainy season lasts about 100 to 140 days, with rainier days in the northern part of Zambia. Malaria transmission occurs throughout the year, but higher prevalence is observed a few months after the rainy season. A higher prevalence of malaria is experienced in the northern part of Zambia(19). Environmental and socioeconomic factors affect the northern part of the Congo River basin. Swampy terrain and equatorial climate promote the breeding and transmission of malaria (20–23).

Children whose households belonged to middle, middle high and highest wealth quantiles were less likely to have malaria. This is so because poor people may not have the income to purchase protective items or replace items such as ITNs, and mosquito repellents and provide proper sleeping spaces that allow poor hanging of the ITN and poor housing structures that might not retain the mosquito chemicals on the wall for a longer time. On the contrary, children from higher wealth quantile might afford items for protection from a mosquito bite and access to healthcare. Studies conducted in Kenya and Malawi found that the wealthy had lower malaria odds than low-income families. This was because wealthy homes could acquire items for protection against mosquito bites (16,24).

Children whose haemoglobin levels were less than 6.9g/dl had increased odds of malaria. Children who are anaemic are susceptible to infections such as malaria because of their low immunity system. The low haemoglobin level may be a sign of prolonged malaria infection. The parasite in blood enters the red blood cells, which multiply and burst the red blood cells and infect more red blood cells, thereby reducing the number of red blood cells and causing anaemia. This is consistent with the study conducted in North-west Ethiopia, where children with anaemia were likely to have malaria infection (16,25).

Children whose houses had damaged or incomplete windows and airbrick screens had higher odds of malaria. Screening of windows and airbricks prevents the entry of mosquitoes into structures. Damaged screens increase the likelihood of mosquitoes entering the houses, thereby biting the human suspects. This intervention at the household level of screening windows and airbricks is effective and lasts as long as the screens are not damaged. Other interventions, such as ITN use and IRS, may supplement this. This is similar to a study conducted in Southwest Ethiopia, which concluded that screening house openings protects against malaria infection (26).

Children whose houses had partially open eaves had increased odds of malaria. Open or partially open eaves allow mosquitoes into the house; hence the occupants have a higher risk of malaria infection. A study in Kenya evaluating the effectiveness of screening house eaves as a potential intervention for reducing indoor vector densities and malaria prevalence showed a low prevalence in houses with closed eaves and a significantly reduced vector density in the Children whose households use water from the well, streams and rivers had higher odds of malaria. The homes may not be very far from these water bodies. These water sources support the breeding of mosquitoes, hence the high transmission of malaria parasites. These people also walk through bushes to fetch water, hence being exposed to mosquito bites. A study conducted in sub-Saharan Africa which analysed 49 surveys (23 DHS, 24 MIS and two others), revealed that unprotected water source was related to a high risk of malaria infection and that children from poor socioeconomic status were mostly affected (28).

House reported not having been sprayed last 12 months increased the odds of malaria in under-five children (29-31). Indoor residual spray chemicals repel and kill mosquitos. This protects the habitants of the house from mosquito bites, hence being protected from malaria infection. This is consistent with a study conducted in Uganda which reported that indoor residual spray was protective and that children whose houses were not sprayed had higher odds of malaria (32).

Studies have shown that ITN use reduces malaria infection. In our study, ITN use was not statistically significant. This could be because there was no evidence that they slept under an ITN; hence the household may decide to report what would be pleasing. It could also be that the ITN had holes due to ageing, hence ineffective. A study conducted in Uganda showed that ITNs were protective (32).

This study will help the government and partners to control and eliminate malaria in Zambia. The focus areas should be vector and parasite interventions which address issues surrounding older children, the population in the rural areas, northern parts of Zambia, the wealth quintile, and infrastructures such as water sources and housing structures. Further, this study may help other researchers to compare findings.

## Conclusion

The study aimed to determine the factors associated with malaria infection in under-five children in Zambia. An increase in age, living in a rural area, and living in northern Zambia was associated with malaria infection. Being in a middle and middle-high-wealth quintile reduced malaria infection in children. Low haemoglobin levels increase the likelihood of malaria. Using a well/stream/river water, windows not screened, and households not sprayed against mosquitoes were associated with malaria. Providing mosquito nets to older children, providing malaria preventive measures to rural communities, implementing integrated malaria interventions in the northern parts of Zambia and conducting IRS may reduce malaria infection. Improving housing structures and providing safe water may reduce the malaria burden in under-five children.

### Limitations

The microscopy dataset and the main MIS dataset could not be merged as there were errors with the barcodes in the main dataset. The dataset could not measure participants’ knowledge because it was not part of it. The study could not address causality because it was a cross-sectional data set.

## Data Availability

All data produced in the present study are available upon reasonable request to the authors

## Authors’ Contributions

LT conceived, designed and implemented the study. CS, NS, MM, and ESH made intellectual input to study design and implementation. NM provided the overall guidance. All authors reviewed the final manuscript and approved it for submission.

## Acknowledgements

The authors acknowledge the ZFETP to be part of the Field Epidemiology Training Program, specifically Dr Jonas Hines, Warren Malambo and Ernest Kateule. Dr Busiku from the Ministry of Health’s National Malaria Elimination Centre, Dr Danny Katongo, Public Health Specialist for Luapula Province. We thank the Centers for Disease Prevention and Control, Zambia National Public Health Institute, Levy Mwanawasa Medical University and the US President’s Malaria Initiative (PMI).

