## Supplementary material for "Factors Associated with Malaria Infection in Under Five Children-Zambia, 2018. A Secondary Analysis of the Malaria Indicator Survey, 2018": Table 1

**Table 1 Background	Characteristics	of the study	Population**

| **Variable** | **n=1,817**  **RDT - (%)** | **n=583**  **RDT + (%)** | **P. Value** |
| --- | --- | --- | --- |
| **Age of children in months** | 1817 (100) | 583 (100) | **<0.0001** |
| Median Age (Kruskal-Wallis test) | 28 | 38 | **<0.0001** |
| **Sex of respondent** |  |  | 0.76 |
| female | 878 (48.3) | 273 (46.8) |  |
| Male | 939 (51.7) | 310 (53.2) |  |
| **Rural/Urban** |  |  | **<0.0001** |
| Urban | 387 21.3) | 21 (3.6) |  |
| Rural | 1430 (78.7) | 562 (96.4) |  |
| **Province** |  |  | **<0.0001** |
| Central | 135 (7.4) | 16 (2.7) |  |
| Copperbelt | 87 (4.8) | 36 (6.2) |  |
| Eastern | 314 (17.3) | 86 (14.8) |  |
| Luapula | 269 (14.8) | 192 (32.8) |  |
| Lusaka | 125 (6.9) | 2 (0.3) |  |
| Muchinga | 124 (6.8) | 56 (9.6) |  |
| North-We | 166 (9.2) | 41 (7.0) |  |
| Northern | 103 (5.6) | 63 (10.8) |  |
| Southern | 168 (9.3) | 2 (0.3) |  |
| Western | 326 (17.9) | 90 (15.4) |  |
| **Wealth quantile** |  |  | **<0.0001** |
| Very low | 301 (16.6) | 201 (34.5) |  |
| Low | 368 (20.3) | 166 (28.5) |  |
| Medium | 419 (23.1) | 123 (21.1) |  |
| High | 381 (21.0) | 85 (14.6) |  |
| Very high | 348 (19.2) | 8 (1.4) |  |
| **Household level of education** |  |  | 0.90 |
| Tertiary | 136 (7.5) | 4 (0.7) |  |
| Non | 363 (20.0) | 154 (26.5) |  |
| Primary | 750 (41.3) | 283 (48.7) |  |
| Secondary | 567 (31.2) | 140 (24.1) |  |
| Tertiary | 136 (7.5) | 4 (0.7) |  |
| **Household source of water** |  |  | **<0.0001** |
| Piped/borehole | 943 (51.9) | 175 (30.1) |  |
| Well/streams | 873 (48.1) | 406 (69.9) |  |
| **Household Toilet** |  |  | **<0.0001** |
| Flushable | 134 (7.4) | 1 (0.2) |  |
| No toilet | 323 (17.8) | 150 (25.8) |  |
| Pit latrine/ VIP/Compost | 1359 (74.8) | 430 (74.0) |  |
| **Type of wall** |  |  | **<0.0001** |
| Finished | 1061 (58.4) | 254 (43.7) |  |
| Natural/rudimentary | 755 (41.6) | 327 (56.3) |  |
| **Roof Material** |  |  | **<0.0001** |
| Finished | 913 (50.3) | 141 (24.3) |  |
| Natural/rudimentary | 903 (49.7) | 440 (75.7) |  |
| **Ceiling** |  |  | **<0.0001** |
| Completely sealed | 54 (3.0) | 1 (0.2) |  |
| Partial | 7 (0.4) | 1 (0.2) |  |
| Non | 1756 (96.6) | 581 (99.7) |  |
| **Windows** |  |  | **<0.0001** |
| Windows/airbricks with screens | 496 (51.2) | 97 (37.0) |  |
| Windows/airbricks screens(holes/incomplete) | 150 (15.5) | 58 (22.1) |  |
| Screens absent | 322 (33.3) | 107 (40.8) |  |
| No window | 848 (46.7) | 1167 (48.7) |  |
| **IRS done** |  |  | 0.24 |
| Yes | 854 (47.0) | 258 (44.3) |  |
| No | 963 (53.0) | 325 (55.7) |  |
| **Eaves** |  |  | **<0.0001** |
| Completely sealed | 782 (43.1) | 199 (34.3) |  |
| Non | 907 (49.9) | 320 (55.1) |  |
| Partially sealed | 127 (7.0) | 62 (10.7) |  |
| **Ill with fever** |  |  | **<0.0001** |
| No | 1506 (82.9) | 372 (63.8) |  |
| Yes | 311 (17.1) | 211 (36.2) |  |
| **Time go out of bed** |  |  | 0.41 |
| Before 22 hours | 1789 (98.3) | 568 (99.8) |  |
| After 22 hours | 10 (1.7) | 1 (0.2) |  |
| **Haemoglobin level** |  |  | **<0.0001** |
| >6.9 | 1750 (99.0) | 537 (95.9) |  |
| <6.9 | 17 (1.0) | 23 (4.1) |  |
| **A.Gambiae%** |  |  | **<0.0001** |
| 5-50 | 1524 (83.9) | 579 (99.3) |  |
| 51-74 | 293 (16.1) | 4 (0.7) |  |
| **A.Fenistus%** |  |  | **<0.0001** |
| 26-50 | 818 (45.0) | 182 (31.2) |  |
| 50.1-89.9 | 269 (14.8) | 104 (17.8) |  |
| >89.9 | 730 (40.2) | 297 (50.9) |  |
| **Rainfall** |  |  | **<0.0001** |
| 148.3-200.0 | 660 (36.3) | 202 (34.6) |  |
| 200.1-250.0 | 1011 (55.6) | 272 (46.7) |  |
| >250.0 | 146 (8.0) | 109 8.7) |  |
