## Supplementary material for "Factors Associated with Malaria Infection in Under Five Children-Zambia, 2018. A Secondary Analysis of the Malaria Indicator Survey, 2018": Table 2

**Table 2 Associations between Outcome and Independent Variables _Full Model**

| **Variable** | **uOR (CI)** | **P-Value** | **aOR (CI)** | **P-Value** |
| --- | --- | --- | --- | --- |
| **Child age in months** | 1.003 (1.002-1.005) | <0.001 | 1.004 (1.003-1.005) | <0.001 |
| **Child gender** |  |  |  |  |
| Female | Ref |  |  |  |
| Male | 1.00 (0.97, 1.03) | 0.99 |  |  |
| **Rural/Urban** |  |  |  |  |
| Urban | Ref |  |  |  |
| Rural | 1.26 (1.20,1.31) | <0.0001 | 1.11 (1.05-1.17) | 0.0003 |
| **Province** |  |  |  |  |
| Central | Ref |  | Ref |  |
| Copperbelt | 1.01(0.95, 1.08) | 0.7 | 1.16 (1.05-1.28) | 0.01 |
| Eastern | 1.09(0.96, 1.23) | 0.18 | 1.08 (0.93-1.26) | 0.32 |
| Luapula | 1.38(1.24, 1.53) | <0.0001 | 1.32 (1.16-1.51) | <0.001 |
| Lusaka | 0.91(0.87, 0.95) | <0.0001 | 1.02 (0.93-1.12) | 0.69 |
| Muchinga | 1.32(1.12, 1.56) | 0.002 | 1.23 (1.06-1.42) | 0.01 |
| Northern | 1.11(1.03, 1.21) | 0.001 | 1.08 (098-1.20) | 0.14 |
| N-Western | 1.34(1.12, 1.60) | 0.001 | 1.34 (1.14-1.59) | 0.001 |
| Southern | 0.92(0.88, 0.96) | 0.0004 | 0.99 (0.91-1.08) | 0.83 |
| Western | 1.12(1.02, 1.23) | 0.01 | 1.06 (0.95-1.19) | 0.29 |
| **Household wealth quantile** |  |  |  |  |
| Lowest | Ref |  | Ref |  |
| Second lowest | 0.89 (0.08, 0.98) | 0.01 | 0.94 (0.88-1.01) | 0.11 |
| Middle | 0.81 (0.74, 0.88) | <0.0001 | 0.89 (0.83-0.95) | 0.001 |
| Middle High | 0.78 (0.71, 0.85) | <0.0001 | 0.91 (0.81-0.99) | 0.04 |
| Highest | 0.69 (0.63, 0.74) | <0.0001 | 0.90 (0.81-1.00) | 0.05 |
| **Head of household education level** |  |  |  |  |
| No education | Ref |  | Ref |  |
| Primary level | 0.96 (0.90 – 1.04) | 0.30 | 0.95 (0.89-1.01) | 0.89 |
| Secondary level | 0.88 (0.82 – 0.94) | <0.0003 | 0.96 (0.91-1.02) | 0.17 |
| Tertiary level | 0.81 (0.75 - 0.88) | <0.001 | 0.93 (0.86 -1.02) | 0.12 |
| **Haemoglobin level** |  |  |  |  |
| >6.9 | Ref |  | Ref |  |
| <6.9 | 1.30 (1.02-1.64) | 0.03 | 1.19 (1.02-1.39) | 0.03 |
| **Time child get out of bed** |  |  |  |  |
| After 04hrs | Ref |  | Ref |  |
| Before 04hrs | 0.87 (0.83-0.90) | <0.001 | 0.94 (0.84-1.04) | 0.22 |
| **Child slept in ITN last night** |  |  |  |  |
| Yes | Ref |  |  |  |
| No | 0.96 (0.90-1.02) | 0.24 |  |  |
| Variable | uOR (CI) | P-Value |  |  |
| **IRS done** |  |  |  |  |
| Yes | Ref |  | Ref |  |
| No | 0.94 (0.88-1.00) | 0.04 | 1.05 (1.01-1.09) | 0.02 |
| **Type of walls of the house** |  |  |  |  |
| Finished walls | Ref |  | Ref |  |
| Rudimentary/natural walls | 1.17 (1.10-1.24) | <0.001 | 1.00 (0.95-1.05) | 0.99 |
| **Type of roof** |  |  |  |  |
| Finished walls | Ref |  | Ref |  |
| Rudimentary/natural walls | 1.23 (1.15-1.31) | <0.001 | 1.01 (0.95-1.08) | 0.68 |
| **Eaves** |  |  |  |  |
| Completely closed | Ref |  | Ref |  |
| Partially open | 1.05 (1.01-1.18) | 0.02 | 0.95 (0.91-0.99) | 0.01 |
| Completely open eaves | 1.09 (0.97-1.22) | 0.15 | 1.02 (0.96-1.08) | 0.59 |
| **Ceiling** |  |  |  |  |
| Completely sealed | Ref |  | Ref |  |
| Partially sealed | 1.00 (0.95-1.05) | 0.9 | 0.89 (0.80-0.99) | 0.03 |
| No ceiling | 1.17 (1.12-1.22) | <0.001 | 1.03 (0.97-1.11) | 0.35 |
| **Windows** |  |  | Ref |  |
| Window/airbrick completely sealed | Ref |  | 1.08 (1.01-1.16) | 0.03 |
| Windows/airbricks screens(holes/incomplete) | 1.07 (0.99-1.16) | 0.08 | 1.02 (0.999-1.08) | 0.4 |
| Screens absent | 1.10 (1.05-1.16) | 0.0003 | 1.02 (0.99-1.07) | 0.21 |
| No window/airbrick | 1.14 (1.08-1.19) | <0.001 |  |  |
| **Household source of water** |  |  | Ref |  |
| Piped/borehole | Ref |  | 1.05 (1.01-1.11) | 0.03 |
| Well/streams/other | 1.18(1.11, 1.25) | <0.0001 |  |  |
| **Type of toilet** |  |  |  |  |
| Flush/Pour flush | Ref |  | Ref |  |
| No Toilet | 1.33 (1.23-1.44) | <0.001 | 1.06 (0.98-1.15) | 0.16 |
| Pit latrine/ VIP/ Compost | 1.18 (1.15-1.22) | <0.001 | 1.02 (0.98-1.05) | 0.34 |
| **Fuel used by household** |  |  |  |  |
| Electricity/gas | Ref |  | Ref |  |
| Charcoal/cooking sticks etc | 1.18 (1.14-1.23) | <0.001 | 1.02 (0.98-1.07) | 0.31 |
| **Provincial vector distribution of A.Fenistus** | 1.003 (1.002-1.004) | <0.001 |  |  |
| **Provincial vector distribution of A. Gambiae** | 0.997 (0.996-0.998) | <0.001) |  |  |
| **District average rainfall** | 1.002 (1.001-1.003) | 0.0003 | 1.00 (1.00-1.00) | 0.31 |

uOR = Unadjusted Odds Ratio, aOR = Adjusted Odds Ratio
