## Supplementary material for "Factors Associated with Malaria Infection in Under Five Children-Zambia, 2018. A Secondary Analysis of the Malaria Indicator Survey, 2018": Table 3

**Table 3 Model Fitness Assessment**

|  | **AIC** | **Loglikelihood** | **Pseudo R2** |
| --- | --- | --- | --- |
| No variable removed | 12401.479 | 116.9439 | 0.255033 |
| Wall removed | 12490.893 | 118.269 | 0.255033 |
| Fuel | 12490.893 | 118.269 | 0.254313 |
| **Roof removed** | **12329.828** | **115.9127** | **0.253888** |
| Eaves removed | 12362.684 | 116.4226 | 0.252153 |
| Window removed | 12365.216 | 116.516 | 0.250389 |
| Rainfall removed | 12376.564 | 116.7464 | 0.250134 |
| Ceiling removed | 12376.564 | 116.7464 | 0.249884 |
| Education removed | 12490.893 | 118.269 | 0.248352 |
