## Supplementary material for "Factors Associated with Malaria Infection in Under Five Children-Zambia, 2018. A Secondary Analysis of the Malaria Indicator Survey, 2018": Table 4

**Table 4 Associations between Outcome and Independent Variables _ Reduced Model**

| **Variable** | **Adjusted OR (CI)** | **P-Value** |
| --- | --- | --- |
| **Child age in months** | **1.003 (1.002 - 1.01)** | **<0.001** |
| **Rural/Urban** |  |  |
| Urban | Ref |  |
| **Rural** | **1.12 (1.06 - 1.18)** | **0.0002** |
| Province |  |  |
| Central | Ref |  |
| **Copperbelt** | **1.14 (1.03 - 1.26)** | **0.01** |
| Eastern | 1.08 (0.93 - 1.25) | 0.31 |
| **Luapula** | **1.35 (1.20 - 1.52)** | **<0.001** |
| Lusaka | 1.04 (0.95 - 1.13) | 0.40 |
| **Muchinga** | **1.25 (1.08 - 1.44)** | **0.004** |
| Northern | 1.10 (1.0001 - 1.21) | 0.05 |
| **N-Western** | **1.34 (1.13 - 1.58)** | **0.001** |
| Southern | 0.97 (0.90 - 1.05) | 0.44 |
| Western | 1.07 (0.97 - 1.18) | 0.20 |
| Household wealth quantile |  |  |
| Lowest | Ref |  |
| Second lowest | 0.93 (0.87 - 1.01) | 0.08 |
| **Middle** | **0.88 (0.82 - 0.94)** | **0.0003** |
| **Middle High** | **0.91 (0.84 - 0.99)** | **0.03** |
| **Highest** | **0.88 (0.79 - 0.97)** | **0.01** |
| **Head of household education level** |  |  |
| Tertiary level | Ref |  |
| No education | 0.95 (0.90 - 1.01) | 0.10 |
| Primary level | 0.96 (0.91 - 1.02) | 0.17 |
| Secondary level | 0.93 (0.86 - 1.02) | 0.10 |
| **Haemoglobin level** |  |  |
| >6.9 | Ref |  |
| **<6.9** | **1.20 (1.02 - 1.42)** | **0.03** |
| **Time child got out of bed** |  |  |
| After 04hrs | Ref |  |
| Before 04hrs | 0.92 (0.84 - 1.02) | 0.11 |
| **Eaves** |  |  |
| Completely closed | Ref |  |
| **Partially open** | **0.95 (0.91-0.99)** | **0.02** |
| Completely open eaves | 1.02 (0.96-1.08) | 0.47 |
| **Ceiling** |  |  |
| Completely sealed | Ref |  |
| **Partially sealed** | **0.89 (0.80 - 0.99)** | **0.03** |
| No ceiling | 1.04 (0.97 - 1.10) | 0.26 |
| **Windows/Airbrick** |  |  |
| Window/airbrick completely sealed | Ref |  |
| Screens absent | 1.03 (0.97 - 1.08) | 0.37 |
| No window/airbrick | 1.02 (0.98 - 1.07) | 0.37 |
| **Household source of water** |  |  |
| Piped/borehole | Ref |  |
| **Well/streams/other** | **1.06 (1.01 - 1.11)** | **0.02** |
| **Type of toilet** |  |  |
| Flush/pour flush | Ref |  |
| No Toilet | 1.07 (0.98 - 1.16) | 0.12 |
| Pit latrine/ VIP/ Compost | 1.03 (0.98 - 1.08) | 0.22 |
| IRS done |  |  |
| Yes | Ref |  |
| **No** | **1.05 (1.01 - 1.10)** | **0.01** |
| District average rainfall | 1.001 (1.00 - 1.001) | 0.31 |
